# Predictors of COVID-19 incidence, mortality, and epidemic growth rate at the country level

**DOI:** 10.1101/2020.05.15.20101097

**Authors:** Nicole Y. Leung, Michelle A. Bulterys, Philip L. Bulterys

## Abstract

**Background:** The burden of the coronavirus disease 2019 (COVID-19) pandemic has been geographically disproportionate. Certain weather factors and population characteristics are thought to drive transmission, but studies examining these factors are limited. We aimed to identify weather, sociodemographic, and geographic drivers of COVID-19 at the global scale using a comprehensive collection of country/territory-level data, and to use discovered associations to estimate the timing of community transmission.

**Methods:** We examined COVID-19 cases and deaths reported up to May 2, 2020 across 205 countries and territories in relation to weather data collected from capital cities for the eight weeks prior to and four weeks after the date of the first reported case, as well as country/territory-level population, geographic, and planetary data. We performed univariable and multivariable regression modeling and odds ratio analyses to investigate associations with COVID-19 cases, deaths, and epidemic growth rate. We also conducted maximum likelihood analysis to estimate the timing of initial community spread.

**Findings:** Lower temperature (p<0.0001), lower humidity (p=0.006), higher altitude (p=0.0080), higher percentage of urban population (p<0.0001), increased air travelers (p=0.00019), and higher prevalence of obesity (p<0.0001) were strong independent predictors of national COVID-19 incidence, mortality, and epidemic growth rate. Temperature at 5–7 weeks before the first reported case best predicted epidemic growth, suggesting that significant community transmission was occurring on average 1–2 months prior to detection.

**Conclusions:** The results of this ecologic analysis demonstrate that global COVID-19 burden and timing of country-level epidemic growth can be predicted by weather and population factors. In particular, we find that cool, dry, and higher altitude environments, as well as more urban and obese populations, may be conducive to more rapid epidemic spread.

## BACKGROUND

Coronavirus disease 2019 (COVID-19) is a newly emerged infectious disease that has caused unprecedented global suffering and mortality since its emergence in late 2019 in Wuhan, China. COVID-19 is caused by the severe acute respiratory syndrome virus coronavirus 2 (SARS-CoV-2), an enveloped single stranded RNA virus that is closely related to SARS coronavirus 1 (SARS-Cov-1) and middle east respiratory syndrome coronavirus (MERS-CoV) (1). SARS-CoV-2 is a zoonotic virus that likely originated in horseshoe bats (*Rhinolophus affinis*), with recent genomic evidence suggesting subsequent passage through an intermediate mammalian host, possibly the heavily trafficked and critically endangered pangolin (mammalian order Pholidota) (2, 3). Following spillover into humans, SARS-CoV-2 displayed remarkably efficient human-to-human spread, with transmission likely mediated primarily by aerosols and respiratory droplets (4, 5). The exponential spread of the virus quickly overwhelmed initial containment efforts, leading to a global pandemic of historic proportions.

Many aspects of COVID-19 epidemiology have yet to be elucidated. For reasons that are not immediately apparent, some countries have experienced significantly higher incidence, mortality, and epidemic growth rate than others. Many factors have been proposed to explain the marked discrepancies in national incidence and mortality following introduction of COVID-19, including weather, demographic, health, and geographic factors (6–8). For instance, it has been proposed that COVID-19, like other droplet-borne respiratory viruses, may spread more efficiently in cold and dry climates (9). Studies in support of this ‘low temperature-low humidity’ theory have focused primarily on incidence and weather data from cities across China, with contradictory findings (10, 11). COVID-19 severity has also been linked to chronic medical conditions, male gender, older age, air pollution, and cigarette smoking (12–15). National case and death counts are also expected to be affected by differences in COVID-19 containment and mitigation strategies, testing capabilities and coverage, health access, and reporting of cases and deaths. Identification of factors that are associated with COVID-19 incidence and mortality and rate of spread on the global scale may shed light on COVID-19 risk factors, transmission, and epidemic dynamics. Therefore, our objective was to test hypotheses for observed differences in global incidence, mortality, and epidemic growth rate by examining country-level COVID-19 case and death data in relation to a suite of publicly available weather, demographic, health, geographic, and planetary data.

## METHODS

### Study design

In this global country-level analysis, we included all cases and deaths reported across 205 countries and territories that have reported at least one COVID-19 case. Daily COVID-19 case and death counts in each country were collected up to May 2, 2020 from the WHO and European Union Centers for Disease Control database (16). We collected the following demographic data for the corresponding countries and territories: median age, population age 0–14 years (%), population age 15–64 years (%), population over 65 years (%), sex ratio, population size (number), population density (pop/km^2^), urban population (%), Gross Domestic Product (GDP) per capita, Human Development Index (HDI), number of airports, and number of air travelers per annum. Sources of demographic data included the CIA World Factbook (for median age, sex ratio, urban population, GDP, number of airports) (17), the World Bank (for age structure, number of airline passengers carried) (18), the International Monetary Fund (for GDP) (19), and the United Nations Human Development Program (for HDI) (20).

We collected the following population-level health data: average body mass index (BMI), obesity (%), diabetic (%), cigarette consumption (annual average per capita), hospital beds (per 100,000 population), physicians (per 100,000 pop), health expenditure (US$ per capita), PM2.5 (µg/m^3^), ozone exposure (ppb), household air pollution (%), Climate Risk Index (2018), and number of COVID-19 tests performed (per 100,000 population). The sources of health data include the World Health Organization (for BMI, health expenditure) (21), the CIA World Factbook (for obesity) (17), the World Bank (for diabetes, hospital beds, physicians) (18), the Tobacco Atlas (for cigarette consumption) (22), the Health Effects Institute (for PM2.5, ozone exposure, and household air pollution) (23), and Germanwatch (for Climate Risk Index) (24). We collected the following climate data from weather stations in each capital city from a global climate data repository, TuTiempo.net (25), which has been used in previous epidemiological studies (26, 27): average temperature (°C), maximum temperature (°C), minimum temperature (°C), atmospheric pressure at sea level (hPA), average relative humidity (%), total rainfall and/or snowmelt (mm), average visibility (km), average wind speed (km/h), total days with snow, total days with thunderstorm, and total days with fog. If the capital city weather station had no data or incomplete data, we selected the largest city closest to the capital city. We collected daily weather data for up to 8 weeks before and four weeks after the date of the first reported case. For countries with a land mass > 1,000,000 km^2^ we collected climate data from the city where the first confirmed case occurred for sensitivity analyses. We also gathered data on population size, population density, COVID-19 testing, and number of days of sunshine from sources that compiled the latest censuses and official estimates or projections.

### Statistical analysis

We used total number of COVID-19 cases, total number of deaths, and cumulative number of cases at 28 days after the first reported case as our main outcomes. The number of cases at four weeks (28 days) was used as a measure of the epidemic growth rate, and enabled normalization of differences in epidemic duration across countries. We averaged (temperature, humidity, atmospheric pressure, visibility, wind speed) or summed (precipitation, days of snow, days of fog, days of thunderstorm) weather variables by week (for sufficient resolution to generate an estimate of the timing of early epidemic spread) and by month prior to the first reported case (for regression modeling). We performed univariable and multivariable negative binomial regression analyses to examine COVID-19 outcomes in relation to weather, demographic, health, and geographic variables as continuous variables. We included variables significant in univariate analyses, as well as those selected *a priori* based on hypotheses regarding increased risk of infection, in multivariable analyses. We also calculated the odds of all outcomes (total cases, total deaths, and cases at 28 days) as categorical variables (total cases <650 or ≥650, total deaths <15 or ≥15, cases at 28 days <100 or ≥100) by stratifying on categories of weather, demographic, health, and geographic variables. Variable cutoffs were set to obtain roughly equal counts across low, middle, and high categories for each variable, and outcome cutoffs were set to obtain roughly equal counts in the low and high outcome categories. Temperature was stratified into five categories, for higher resolution, while variables with high ‘0’ or ‘1’ count data were stratified into two categories instead of three.

We fit negative binomial regression models to examine the hypothetical timing of COVID-19 exposure. This was achieved by subtracting a range of exposure-to-presentation delays from the day of first reported case and examining likelihood scores for models fit with the different delay durations. Maximum likelihood estimates were generated by calculating the negative log-likelihood (NLL) from the Akaike information criterion (AIC) of a given model, as previously described (26). All log-likelihood scores within 1.92 units of the maximum score were considered to be within the 95% confidence interval (28). All statistical analyses were performed using R (version 4.0.0). Regressions were performed using the glm.nb function in the MASS package with a log-link function. Given overdispersion of count data, a likelihood ratio test was used to determine that the negative binomial regression model was required instead of a standard Poisson model. The Mann-Whitney test was used for country profile comparisons.

## Results

### COVID-19 burden by country and continent

As of May 2, 2020, 3,306,882 COVID-19 cases and 238,424 COVID-19 deaths were reported globally (overall case fatality rate: 7.2%). We examined cumulative cases, deaths, and epidemic growth rates (cumulative cases in the first 28 days after the first reported case) reported in 205 countries and territories reporting at least one COVID-19 case (Supplementary Table 1). The epidemic growth rate varied greatly by country/territory: in the first 28 days after the first reported case, the cumulative number of cases ranged from 1 to 34,109 cases (median: 91 cases) (Figure 1A). Of total cases in the first 28 days, 45.7% occurred in Asia, 36.1% in Europe, 7.7% in South America, 5.3% in Africa, 4.9% in North America., and 0.4% in Oceania (Figure 1B). The number of deaths in the first 28 days was also highly variable and largely reflected case distributions (Figures 1C and 1D). The total number of cases and deaths up to May 2, 2020 also varied greatly (see Supplementary Figures 1A and 1C), with cases ranging from 3 to 1,103,781 (median: 644) and deaths ranging from 0 to 65,068 (median: 13). Of total cases, 36.9% occurred in Europe, 36.3% in North America, 19.6% in Asia, 5.7% in South America, 1.2% in Africa, and 0.3% in Oceania (Supplementary Figure 1B). Of total deaths, 57.0% occurred in Europe, 29.9% in North America, 8.5% in Asia, 3.9% in South America, 0.7% of deaths in Africa, and 0.1% in Oceania (Supplementary Figure 1D). Countries/territories reported their first case in the months of December (n=1), January (n=21), February (n=36), March (n=138), and April (n=9). Epidemic duration ranged from 23 to 124 days (median: 54 days), although countries did not have the same amount of time in observation.

**Figure 1.**
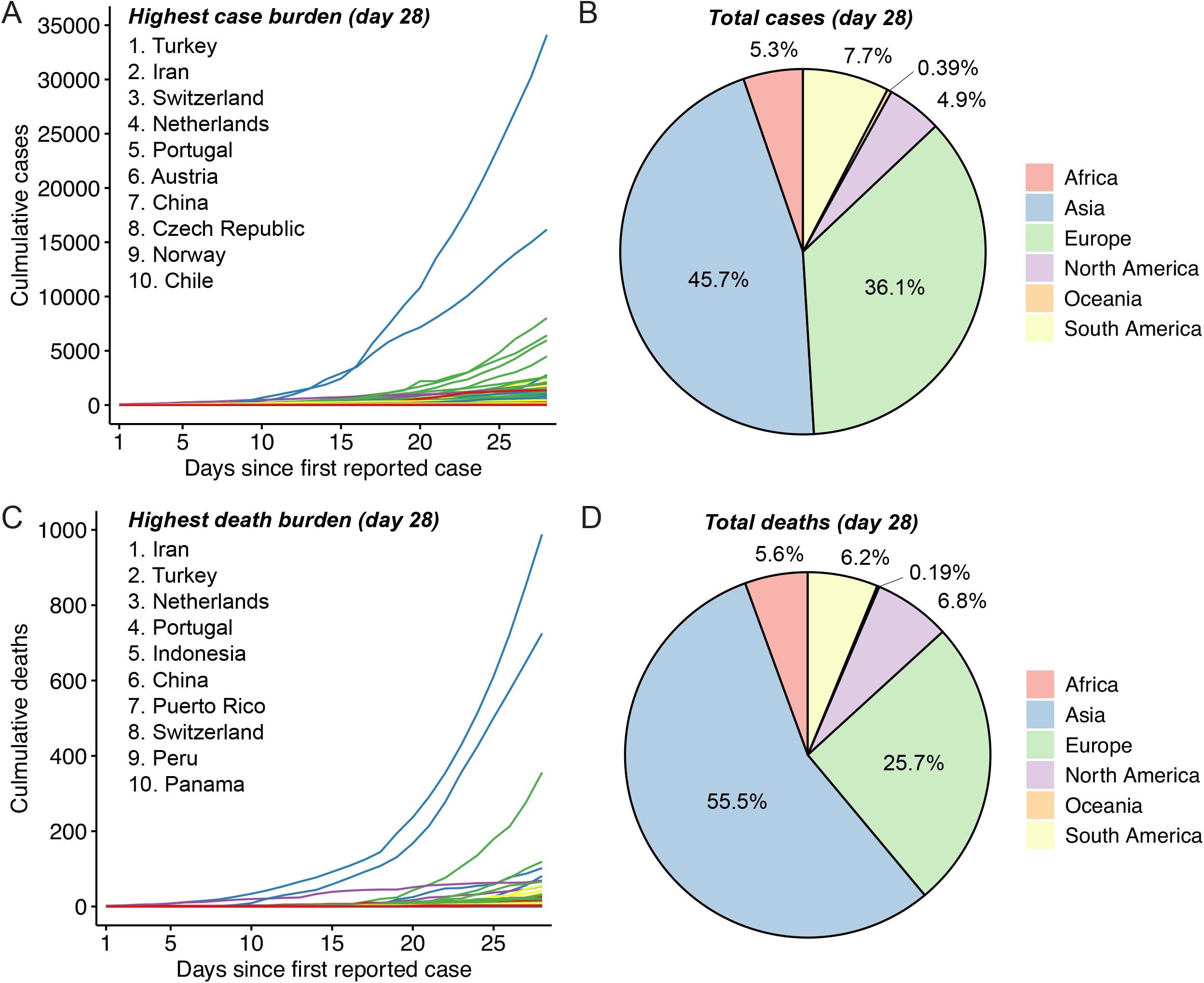
COVID-19 incidence and mortality by country and continent. (A) Cumulative COVID-19 cases by country up to 28 days since first reported case. Listed are countries with the highest case burden 28 days after first reported case. Each country is colored by continent: Africa (red), Asia (blue), Europe (green), North America (purple), Oceania (orange), and South America (yellow). (B) Total COVID-19 cases by continent at 28 days as of May 2, 2020 (percentage of total; number of cases): Asia (45.69%; 62,771), Europe (36.09%; 49,588), South America (7.65%; 10,515), Africa (5.28%; 7,254), North America (4.90%; 6,728), and Oceania (0.39%; 542). (C) Cumulative COVID-19 deaths by country up to 28 days since first reported case. Listed are countries with the highest total death burden 28 days after first reported case. Each country is colored by continent. (D) Total COVID-19 deaths by continent at 28 days as of May 2, 2020 (percentage of total; number of deaths): Asia (55.50%; 1,993), Europe (25.70%; 923), North America (6.82%; 245), South America (6.21%; 223), Africa (5.57%; 200), and Oceania (0.19%; 7).

### COVID-19 and country weather factors

Daily weather data for a 12-week period surrounding the date of first reported case (8 weeks prior to four weeks after) were extracted for the capital city of each of 188 countries/territories with available weather data and analyzed in relation to COVID-19 outcomes (total cases, total deaths, and epidemic growth rate). In univariable regression analyses, all examined outcomes were strongly associated with lower temperature (average, minimum, maximum) and lower relative humidity (Table 1 and Figure 2A-C), as well as lower number of days with thunderstorm and lower number of sunshine days (Table 1 and Supplementary Figure 2). In multivariable analysis adjusting for temperature, median age, and average BMI, the only weather variables that remained significantly associated with epidemic growth rate were lower average temperature (p<0.0001), lower minimum temperature (p<0.0001), lower maximum temperature (p<0.0001) and lower relative humidity (p = 0.0055) (Table 1). When examined by temperature, more rapid epidemic growth was evident in countries with average temperatures <15°C (Figure 2D). Countries with temperatures between 6–15°C showed the highest average epidemic growth rate (Figure 2D). Figures 3A and 3B provide a cartographic representation of these data, showing the average temperature in the four weeks preceding the first case in each country, and the case counts at 28 days and total case counts, respectively. Weather variable findings were corroborated when examined in odds ratio (OR) analyses. Countries with low temperature (≤15°C) were at markedly increased odds of having increased total cases, increased total deaths, and rapid epidemic growth relative to countries with high temperature (≥27°C) (Table 2). The odds of rapid epidemic growth were greatest for countries with temperatures between 6 and 15°C (OR: 8.0, 95% CI: 2.9–22.3), followed by countries with temperatures <6°C (OR: 3.4, 95% CI: 1.4–8.6). Countries that experienced at least one day of snow or fog were at significantly increased risk of COVID-19 cases and deaths, while countries with more hours of sunshine and greater visibility were relatively protected (Supplementary Table 2). Rain, humidity, wind, and thunderstorms did not alter the odds of having high case and death counts. For countries with a landmass greater than 1,000,000 km^2^ (n=29), weather data were also collected from the city where the first COVID-19 case was reported, for sensitivity analysis. For seven countries, this city was different from the capital city, and all univariable and multivariable regression results remained robust to this change.

**Figure 2.**
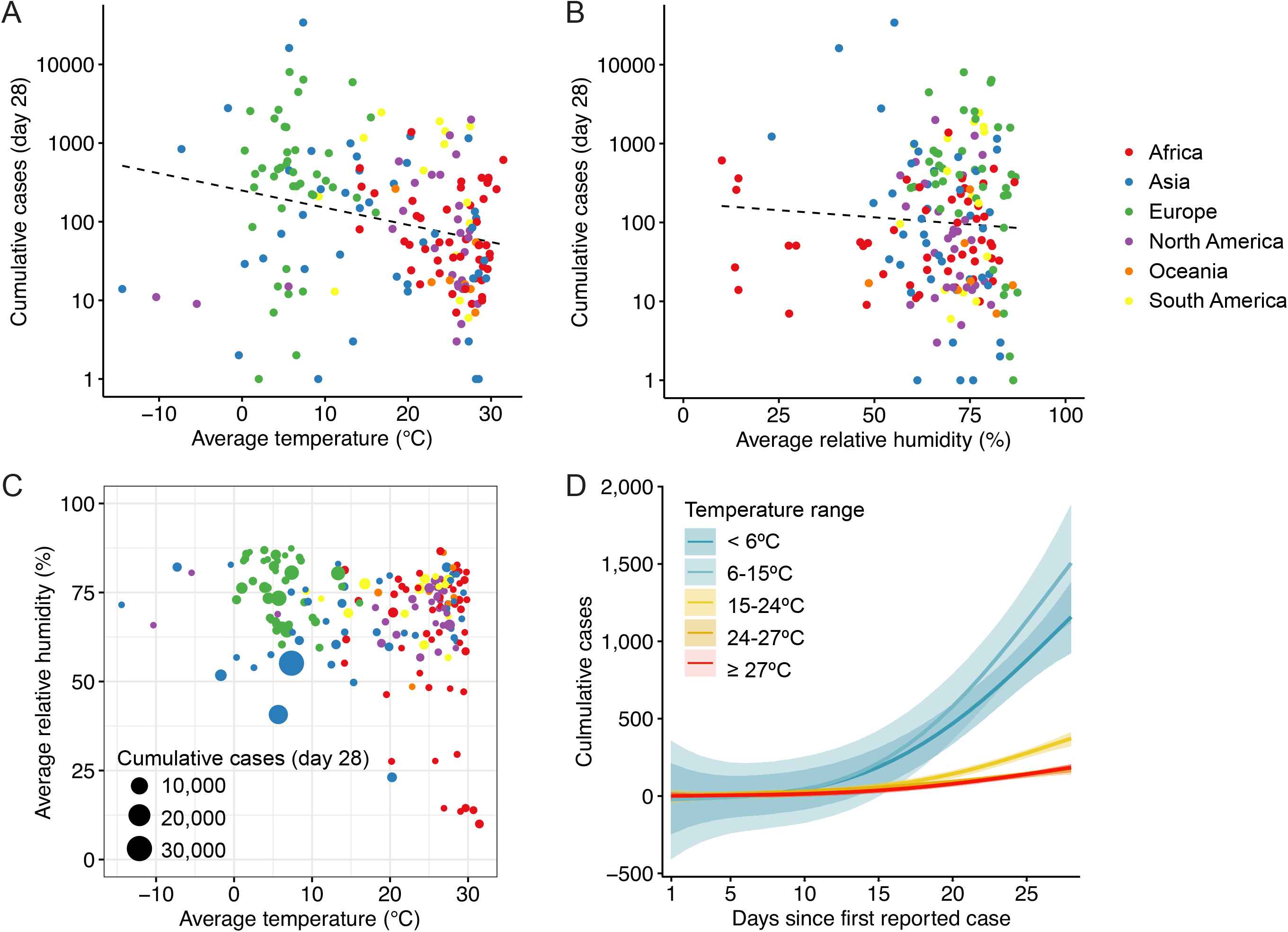
COVID-19 cases in relation to temperature and humidity. (A-B) Cumulative COVID-19 cases by country at 28 days since first reported case in relation to (A) average temperature and (B) average relative humidity. Average temperature represents the mean of average daily temperatures over the four weeks preceding the first reported case. Average relative humidity represents the mean of average daily relative humidity over the four weeks preceding the first reported case. The regression line is fit using a linear model (dashed). Each country is colored by continent. (C) Cumulative COVID-19 cases at 28 days since first reported case by average temperature and average relative humidity. Each country is colored by continent and the number of cases in each country is represented by the size of the data point. (D) Cumulative COVID-19 cases up to 28 days since first reported case for countries stratified into five groups by average temperature: <6°C (blue), between 6-15°C (light blue), between 15-24°C (yellow), between 24-27°C (orange), and =27°C (red). The shading represents 95% confidence intervals.

**Table 1:**
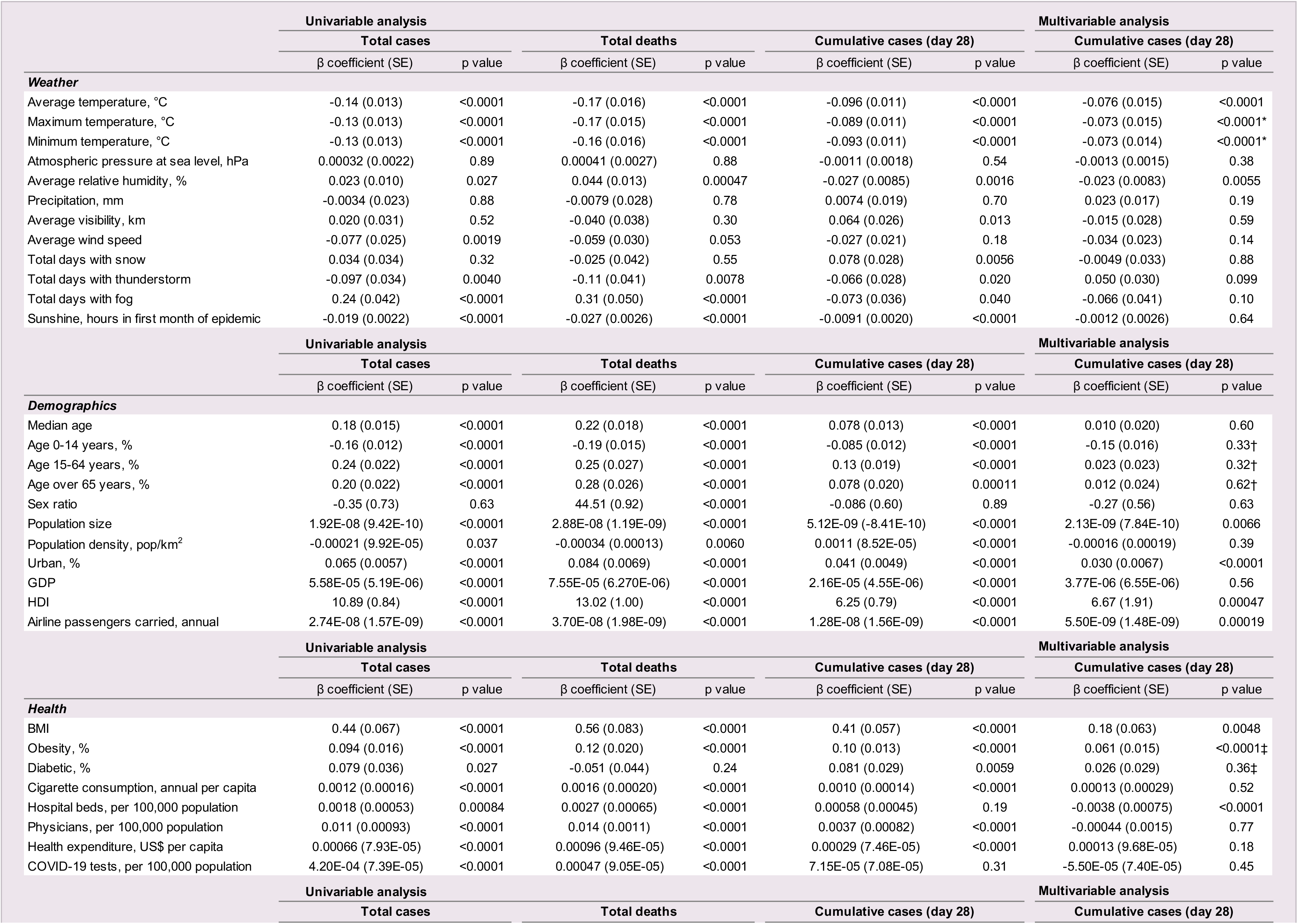
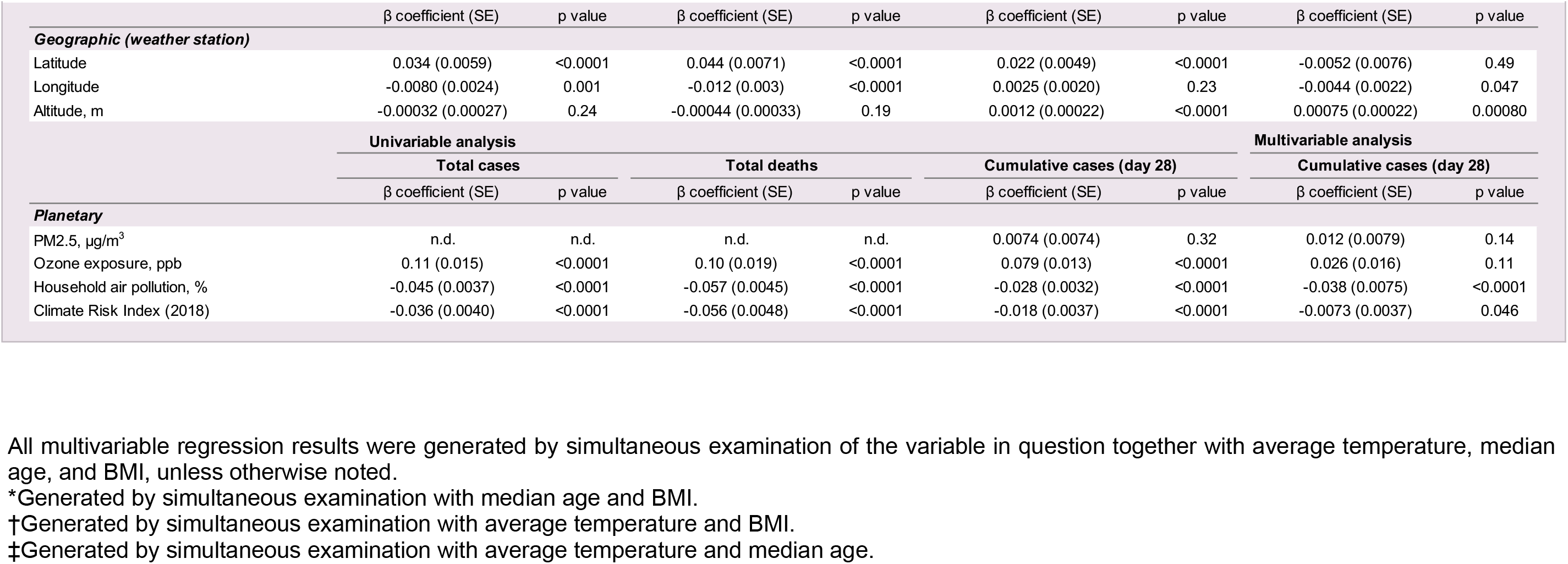
Univariable and multivariable associations between COVID-19 outcomes (cases, deaths, and epidemic growth rate) and weather, demographic, health, geographic, and planetary factors.

### COVID-19 and demographic characteristics at the country level

In univariable regression analyses (Table 1), COVID-19 total cases, total deaths, and epidemic growth rate were strongly associated with older overall median age, lower percentage of population between 0–14 years of age, higher percentage of population over 65 years of age, increased population size, increased population density, increased urban population, increased GDP, increased HDI, and increased number of annual airline passengers. Sex ratio was not associated with cases at 28 days or total cases, but was associated with total deaths (p<0.0001) in univariable regression analysis. In multivariable models (which adjusted simultaneously for temperature, median age, and BMI), increased population size, increased urban population, and higher number of air travelers remained significantly associated with epidemic growth rate (Table 1). Median age was no longer significant when adjusting for temperature and BMI. Countries with increased population size, urban percentage, higher HDI, and annual airline passengers also had greater risk of cases and deaths (Table 2). Sex ratio was not associated with increased deaths in the odds ratio analysis (OR: 0.83, 95% CI: 0.41–1.7) (Supplementary Table 2).

**Table 2:** Odds of COVID-19 outcomes (cases, deaths, and epidemic growth rate) by weather, demographic, health, geographic, and planetary factors.

|  |  | Total cases |  |  | Total deaths |  |  | Cumulative cases (day 28) |  |  |
| --- | --- | --- | --- | --- | --- | --- | --- | --- | --- | --- |
|  |  | <650 | ≥650 | Odds Ratio (95% CI) | <15 | ≥15 | Odds Ratio (95% CI) | <100 | ≥100 | Odds Ratio (95% CI) |
| **Weather** | **Stratification** |  |  |  |  |  |  |  |  |  |
|  | Average temperature, °C |  |  |  |  |  |  |  |  |  |
|  | ≥27 | 33 | 14 | 1 (ref) | 31 | 16 | 1 (ref) | 32 | 14 | 1 (ref) |
|  | ≥24 to <27 | 25 | 6 | 0.57 (0.19-1.68) | 25 | 6 | 0.47 (0.16-1.36) | 22 | 9 | 0.94 (0.34-2.54) |
|  | ≥15 to <24 | 18 | 18 | 2.36 (0.78-7.12) | 18 | 18 | 1.94 (0.64-5.85) | 16 | 20 | 2.86 (1.03-7.9.00) |
|  | ≥6 to <15 | 9 | 27 | 7.07 (2.61-19.18) | 8 | 28 | 6.78 (2.44-18.84) | 8 | 28 | 8.00 (2.87-22.29) |
|  | <6 | 3 | 32 | 25.14 (6.59-95.87) | 5 | 30 | 11.63 (3.78-35.72) | 14 | 21 | 3.43 (1.36-8.63) |
| Maximum temperature, °C | ≥30 | 45 | 19 | 1 (ref) | 43 | 21 | 1 (ref) | 43 | 20 | 1 (ref) |
|  | ≥17 to <30 | 33 | 28 | 2.01 (0.96-4.19) | 33 | 28 | 1.74 (0.84-3.59) | 30 | 31 | 2.22 (1.07-4.61) |
|  | <17 | 10 | 50 | 11.84 (4.99-28.13) | 11 | 49 | 9.12 (3.95-21.06) | 19 | 41 | 4.64 (2.17-9.92) |
| Minimum temperature, °C | ≥21 | 48 | 18 | 1 (ref) | 47 | 19 | 1 (ref) | 45 | 20 | 1 (ref) |
|  | ≥8 to <21 | 30 | 28 | 2.49 (1.18-5.26) | 29 | 29 | 2.47 (1.18-5.19) | 28 | 30 | 2.41 (1.15-5.04) |
|  | <8 | 10 | 51 | 13.60 (5.71-32.39) | 11 | 50 | 11.24 (4.84-26.11) | 19 | 42 | 4.97 (2.34-10.59) |
| Average relative humidity, % | ≥76 | 27 | 35 | 1 (ref) | 22 | 40 | 1 (ref) | 29 | 32 | 1 (ref) |
|  | ≥66 to <76 | 35 | 26 | 0.57 (0.28-1.17) | 36 | 25 | 0.38 (0.18-0.79) | 33 | 28 | 0.77 (0.38-1.57) |
|  | <66 | 26 | 35 | 1.04 (0.51-2.12) | 29 | 32 | 0.61 (0.29-1.25) | 30 | 31 | 0.94 (0.46-1.91) |
|  |  | <650 | ≥650 | Odds Ratio (95% CI) | <15 | ≥15 | Odds Ratio (95% CI) | <100 | ≥100 | Odds Ratio (95% CI) |
| **Demographics** | **Stratification** |  |  |  |  |  |  |  |  |  |
|  | Population size |  |  |  |  |  |  |  |  |  |
|  | <2400000 | 55 | 11 | 1 (ref) | 55 | 11 | 1 (ref) | 42 | 23 | 1 (ref) |
|  | ≥2400000 to <16000000 | 28 | 41 | 7.32 (3.27-16.40) | 30 | 39 | 6.5 (2.91-14.52) | 29 | 39 | 2.46 (1.22-4.94) |
|  | ≥16000000 | 22 | 48 | 10.91 (4.80-24.79) | 18 | 52 | 14.44 (6.23-33.47) | 32 | 37 | 2.11 (1.05-4.23) |
|  | Urban, % |  |  |  |  |  |  |  |  |  |
|  | <50 | 50 | 14 | 1 (ref) | 46 | 18 | 1 (ref) | 44 | 18 | 1 (ref) |
|  | ≥50 to <75 | 29 | 41 | 5.05 (2.36-10.80) | 27 | 43 | 4.07 (1.97-8.42) | 25 | 44 | 4.30 (2.06-8.98) |
|  | ≥75 | 25 | 44 | 6.29 (2.91-13.57) | 29 | 40 | 3.52 (1.71-7.28) | 33 | 36 | 2.67 (1.29-5.50) |
|  | HDI |  |  |  |  |  |  |  |  |  |
|  | <0.66 | 45 | 13 | 1 (ref) | 40 | 18 | 1 (ref) | 38 | 17 | 1 (ref) |
|  | ≥0.66 to <0.805 | 25 | 34 | 4.71 (2.11-10.53) | 29 | 30 | 2.30 (1.08-4.89) | 24 | 35 | 3.26 (1.51-7.06) |
|  | ≥0.805 | 7 | 51 | 25.22 (9.25-68.73) | 10 | 48 | 10.67 (4.43-25.71) | 20 | 38 | 4.25 (1.93-9.34) |
|  | Airline passengers carried, annual |  |  |  |  |  |  |  |  |  |
|  | <700000 | 38 | 11 | 1 (ref) | 33 | 16 | 1 (ref) | 31 | 17 | 1 (ref) |
|  | ≥700000 to <7000000 | 21 | 32 | 5.26 (2.21-12.54) | 24 | 29 | 2.49 (1.11-5.58) | 16 | 37 | 4.22 (1.83-9.70) |
|  | ≥7000000 | 2 | 50 | 86.36 (18.06-412.88) | 5 | 47 | 19.39 (6.46-58.15) | 23 | 29 | 2.30 (1.03-5.15) |
|  |  | <650 | ≥650 | Odds Ratio (95% CI) | <15 | ≥15 | Odds Ratio (95% CI) | <100 | ≥100 | Odds Ratio (95% CI) |
| **Health** | **Stratification** |  |  |  |  |  |  |  |  |  |
| BMI | <25 | 39 | 20 | 1 (ref) | 36 | 23 | 1 (ref) | 41 | 17 | 1 (ref) |
|  | ≥25 to <26.7 | 14 | 43 | 5.99 (2.67-13.45) | 15 | 42 | 4.38 (1.99-9.64) | 16 | 39 | 5.88 (2.61-13.23) |
|  | ≥26.7 | 23 | 35 | 2.97 (1.40-6.30) | 26 | 32 | 1.93 (0.92-4.02) | 25 | 33 | 3.18 (1.48-6.86) |
| Obesity, % | <12 | 38 | 19 | 1 (ref) | 33 | 24 | 1 (ref) | 39 | 17 | 1 (ref) |
|  | ≥12 to <23 | 21 | 37 | 3.52 (1.63-7.60) | 24 | 34 | 1.95 (0.93-4.09) | 22 | 34 | 3.55 (1.62-7.75) |
|  | ≥23 | 17 | 42 | 4.94 (2.25-10.86) | 20 | 39 | 2.68 (1.26-5.69) | 21 | 38 | 4.15 (1.90-9.06) |
| Hospital beds, per 100,000 population | <150 | 38 | 21 | 1 (ref) | 34 | 25 | 1 (ref) | 32 | 26 | 1 (ref) |
|  | ≥150 to <340 | 27 | 35 | 2.35 (1.13-4.88) | 28 | 34 | 1.65 (0.80-3.39) | 31 | 30 | 1.19 (0.58-2.45) |
|  | ≥340 | 19 | 43 | 4.10 (1.92-8.74) | 21 | 41 | 2.66 (1.27-5.55) | 25 | 37 | 1.82 (0.88-3.76) |
|  |  | Total cases |  |  | Total deaths |  |  | Cumulative cases (day 28) |  |  |
|  |  | <650 | ≥650 | Odds Ratio (95% CI) | <15 | ≥15 | Odds Ratio (95% CI) | <100 | ≥100 | Odds Ratio (95% CI) |
| **Geographic (weather station)** | **Stratification** |  |  |  |  |  |  |  |  |  |
| Longitude | <-5 | 37 | 23 | 1 (ref) | 37 | 23 | 1 (ref) | 32 | 28 | 1 (ref) |
|  | ≥-5 to <30 | 21 | 39 | 2.99 (1.42-6.28) | 17 | 43 | 4.07 (1.89-8.75) | 22 | 37 | 1.92 (0.92-4.00) |
|  | ≥30 | 30 | 35 | 1.88 (0.92-3.83) | 33 | 32 | 1.56 (0.77-3.18) | 38 | 27 | 0.81 (0.40-1.65) |
| Altitude, m | <20 | 33 | 27 | 1 (ref) | 35 | 25 | 1 (ref) | 37 | 22 | 1 (ref) |
|  | ≥20 to <120 | 27 | 34 | 1.54 (0.75-3.15) | 29 | 32 | 1.54 (0.75-3.17) | 29 | 32 | 1.86 (0.90-3.85) |
|  | ≥120 | 28 | 36 | 1.57 (0.77-3.19) | 23 | 41 | 2.50 (1.21-5.15) | 26 | 38 | 2.46 (1.19-5.08) |
|  |  | Total cases |  |  | Total deaths |  |  | Cumulative cases (day 28) |  |  |
|  |  | <650 | ≥650 | Odds Ratio (95% CI) | <15 | ≥15 | Odds Ratio (95% CI) | <100 | ≥100 | Odds Ratio (95% CI) |
| **Planetary** | **Stratification** |  |  |  |  |  |  |  |  |  |
| PM2.5, µg/m<sup>3</sup> | <50 | 80 | 85 | 1 (ref) | 79 | 86 | 1 (ref) | 79 | 83 | 1 (ref) |
|  | ≥50 | 2 | 14 | 6.59 (1.45-29.91) | 4 | 12 | 2.76 (0.85-8.90) | 7 | 9 | 1.22 (0.43-3.44) |
| Household air pollution, % | <1.5 | 12 | 46 | 1 (ref) | 15 | 43 | 1 (ref) | 23 | 35 | 1 (ref) |
|  | ≥1.5 to <35 | 23 | 36 | 0.41 (0.18-0.93) | 27 | 32 | 0.41 (0.19-0.90) | 21 | 37 | 1.16 (0.55-2.45) |
|  | ≥35 | 47 | 17 | 0.090 (0.040-0.22) | 41 | 23 | 0.20 (0.090-0.43) | 42 | 20 | 0.31 (0.15-0.66) |
| Climate Risk Index (2018) | <65 | 21 | 35 | 1 (ref) | 21 | 35 | 1 (ref) | 26 | 29 | 1 (ref) |
|  | ≥65 to <105 | 19 | 39 | 1.23 (0.57-2.66) | 18 | 40 | 1.33 (0.61-2.90) | 21 | 37 | 1.58 (0.74-3.35) |
|  | ≥105 | 34 | 24 | 0.42 (0.20-0.90) | 37 | 21 | 0.34 (0.16-0.73) | 34 | 23 | 0.61 (0.29-1.28) |

### COVID-19 and health factors at the country level

In univariable regression analyses, epidemic growth rate, cases, and deaths were strongly associated with BMI, high prevalence of obesity, diabetes, cigarette consumption, number of physicians, and health expenditure (Table 1 and Supplementary Figure 2). Number of COVID-19 tests performed was positively associated with total cases (p<0.0001) and total deaths (p<0.0001), but not with epidemic growth rate (p = 0.31). After adjusting for temperature and median age, BMI (p = 0.005) and obesity (p<0.0001) remained positively associated with epidemic growth rate. When adjusting for temperature, median age, and BMI, the number of hospital beds was negatively associated with epidemic growth rate. In odds ratio analyses (Table 2 and Supplementary Table 2), countries with high BMI and high prevalence of obesity, high cigarette consumption, high number of physicians, and high health expenditure were at significantly greater odds of having increased epidemic growth rate. COVID-19 mortality was associated with increased BMI (OR: 4.4, 95% CI: 2.0–9.6), obesity (OR: 2.7, 95% CI: 1.3–5.7), cigarette consumption (OR: 9.5, 95% CI: 3.8–23.8), number of hospital beds (OR: 2.7, 95% CI: 1.3–5.6), number of physicians (OR: 11.0, 95% CI: 4.6–26.3), health expenditure (OR: 8.2, 95% CI: 3.5–19.0), and number of COVID-19 tests performed (OR: 11.0, 95% CI: 2.8–43.1) (Table 2 and Supplementary Table 2).

### COVID-19 and geographic/planetary factors at the country level

Latitude, longitude, and altitude data were collected for each country/territory and analyzed in relation to COVID-19 incidence and mortality. More northern latitude (relative to the equator) was significantly positively associated with total cases (p<0.0001), total deaths (p<0.0001), and epidemic growth rate (p<0.0001) in univariable analysis; however, when examined together with temperature, BMI, and median age in a multivariable model, it was no longer significantly associated (p = 0.49) (Table 1 and Supplementary Figure 3). More eastern longitude (relative to the Prime Meridian) was significantly associated with total cases (p = 0.001) and total deaths (p<0.0001), but not epidemic growth rate (p = 0.23), in univariable analyses. In multivariable analysis, the association between longitude and epidemic growth rate was borderline significant (p = 0.047). Higher altitude was significantly associated with increased epidemic growth rate (p<0.0001) and total deaths (0.0003) in univariable analysis, and remained significantly associated in multivariable analysis (p = 0.0008). Among all planetary factors examined, ozone exposure (p<0.0001) was found to be positively associated with cases at 28 days, total cases, and total deaths, while household air pollution and climate risk index were both negatively associated with cases, in univariable analysis. In multivariable analysis, household air pollution (p<0.0001) and climate risk index (p = 0.046) remained significantly negatively associated with epidemic growth rate, while ozone exposure did not (p = 0.11). In odds ratio analyses, high household air pollution (OR: 0.2, 95% CI: 0.09–0.43) and high climate risk index (OR: 0.34, 95% CI: 0.16–0.73) were associated with reduced odds of rapid epidemic growth, while high ozone exposure was associated with increased odds of high epidemic growth rate (OR: 2.8, 95% CI 1.3–6.0) (Table 2 and Supplementary Table 2).

**Figure 3.**
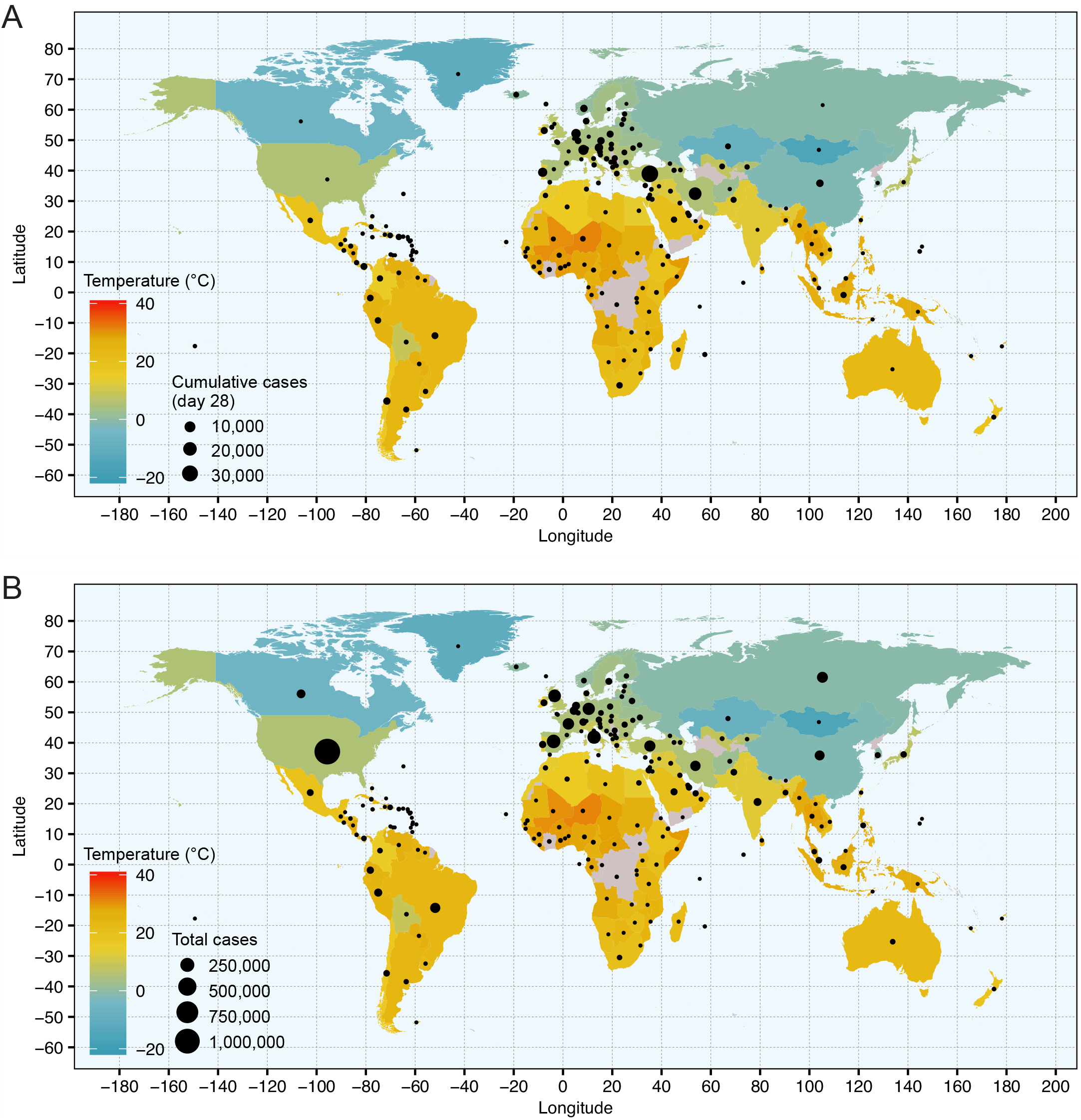
World maps of COVID-19 cases and temperature. World maps showing (A) COVID-19 case counts by country at 28 days since the first reported case and (B) total COVID-19 cases since first reported case up to May 2, 2020. The number of cases is represented by the size of the data point and each country is colored by the mean daily temperature in the capital city over the four weeks preceding the first reported case.

### Profile of highly COVID-19 susceptible and non-susceptible countries

Table 3 compares the profiles of countries with the highest epidemic growth rates (top 20%; n=40) and lowest epidemic growth rates (bottom 20%; n=42). Countries in the top 20% had a median of 1241 cases while countries in the bottom 20% had a median of 9.5 cases at 28 days after the first case. Compared to countries in the bottom 20%, countries in the top 20% had lower temperature (median: 10.4^°^C vs 26.0^°^C), lower relative humidity (69.3% vs 72.2%), higher altitude (106m vs 37m), older median age (35.5 vs. 33.3), increased urban population (74.4% vs 61.3%), higher GDP ($29,441 vs. $15,124), higher BMI (26.3 vs. 25.9), and increased cigarette consumption (1204.3 vs. 635.1 annual per capita).

**Table 3:** Profile of countries with the highest (top 20%) and lowest (bottom 20%) epidemic growth rates.

| ***Factor*** | **Top 20%** | **Bottom 20%** | p value |
| --- | --- | --- | --- |
|  | Median (±SEM) | Median (±SEM) |  |
| Total cases at day 28 | 1,241 (914) | 9.5 (0.7) | <0.0001 |
| Average temperature, °C | 10.4 (1.6) | 26.0 (2.2) | 0.021 |
| Average relative humidity, % | 69.3 (2.5) | 72.2 (2.5) | 0.273 |
| Altitude, m | 106.0 (96.8) | 36.5 (45.2) | 0.017 |
| Age 0-14 years, % | 20.2 (1.2) | 26.3 (1.7) | 0.022 |
| Age over 65 years, % | 11.2 (1.0) | 6.9 (1.2) | 0.061 |
| Obesity, % | 22.5 (1.1) | 21.0 (1.7) | 0.048 |
| Urban, % | 74.4 (2.8) | 61.3 (4.5) | 0.120 |
| GDP | 29,441 (4,552) | 15,124 (3,794) | 0.039 |
| HDI | 0.817 (0.018) | 0.735 (0.030) | 0.023 |
| Airline passengers carried, annual | 15,728,390 (14,580,248) | 8,483,180 (35,359,452) | 0.420 |
| Cigarette consumption, annual per capita | 1,204 (172) | 635 (126) | 0.033 |
| Health expenditure, US\$ per capita | 1,492 (314) | 573 (430) | 0.020 |
| COVID-19 tests, per 100,000 population | 1,148 (474) | 1,771 (768) | >0.9999 |

Countries were categorized in the top or bottom 20% for epidemic growth rate according to cumulative case counts at 28 days after the first reported case. 40 countries were included in the top 20%, and 42 countries were included in the bottom 20% (two additional countries were included in the bottom category as they had the same case count at day 28). Three countries/territories (São Tomé and Príncipe, South Sudan, and Yemen) had not reached 28 days after the first reported case as of May 2, 2020 and were omitted. SEM represents the standard error of the median. P values for comparisons were generated using the Mann-Whitney test.

### Maximum likelihood estimation of epidemic start date

The strong association between temperature and COVID-19 cases allowed us to generate a conditional estimate of the average epidemic start date, relative to the date of first reported case. Temperature was selected because it was the strongest weather predictor of COVID-19 cases in regression analyses. Comparison of the countries with highest vs. lowest epidemic growth rate with the countries with lowest epidemic growth rate revealed significantly lower temperatures among the countries with high case counts (Figure 4A). In addition, for countries with highest epidemic growth rate in the first 28 days, temperatures were lowest 5–7 weeks prior to the first reported case. We used a maximum likelihood approach to generate an estimate of the epidemic start date, conditional on the validity of the association with temperature. We incorporated hypothetical exposure-to-presentation delays in our regression model linking case counts at an intermediate epidemic time point (six weeks) to temperature and compared the goodness-of-fit of these models for delays of 0–8 weeks. The likelihood scores incorporating these delays indicate that the temperature 5–7 weeks prior to the first reported case best predicted the epidemic size at six weeks (Figure 4B), reflecting average temperature patterns (Figure 4A). This suggests that community transmission was occurring on average 1–2 months before the first laboratory-confirmed case.

**Figure 4.**
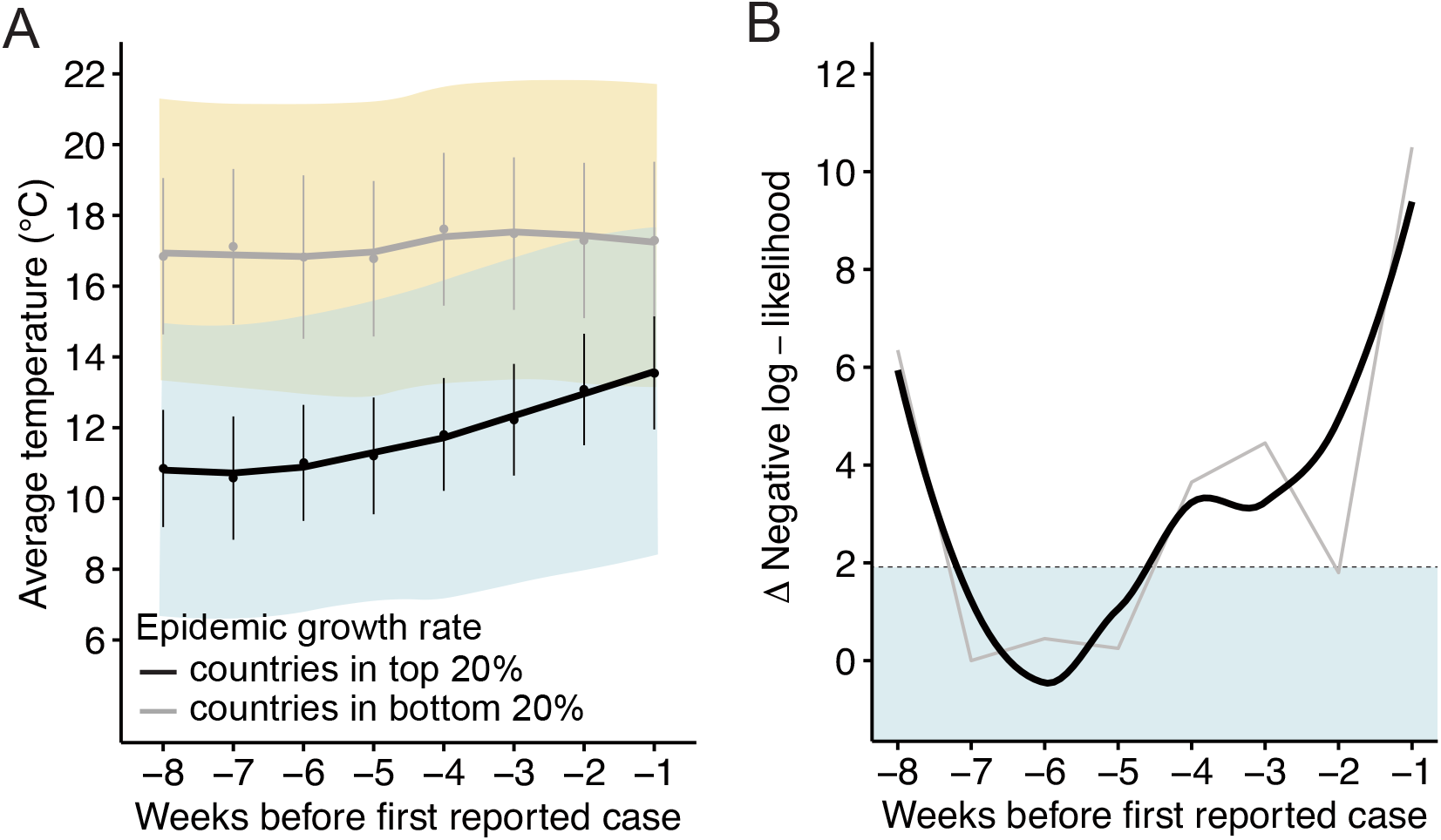
Maximum likelihood estimation of the timing of early epidemic. (A) Weekly average temperatures up to eight weeks preceding the first reported case of countries stratified into two groups according to epidemic growth rate in the first 28 days: top 20%; i.e. highest epidemic growth rates (black; 40 countries) and bottom 20%; i.e. lowest epidemic growth rates (gray; 42 countries). The regression lines are fit using the loess method. The error bars represent ± standard error of the mean (SEM). The shaded regions represent the average minimum-maximum range of temperatures for countries in the top 20% (blue) and bottom 20% (yellow), with the overlapping region shaded green. (B) Negative log-likelihood values from negative binomial regression of cumulative cases at 28 days and weekly average temperatures up to eight weeks preceding the first reported case. The regression line is fit using the loess method (black). The dashed line corresponds to a difference of 1.92 log-likelihood units from the optimum value; points beneath this line fall within the 95% confidence interval.

## DISCUSSION

The goal of this study was to identify weather, demographic, health, geographic, and planetary factors associated with national COVID-19 epidemic growth rate, incidence, and mortality. We included all cases and deaths reported globally from the date of the first reported case up until May 2, 2020, and examined global weather data contemporaneous to the first reported case in each country, as well as a comprehensive suite of demographic, health, geographic, and planetary factors. We identified lower temperature, lower relative humidity, higher altitude, high prevalence of obesity, and higher number of air travelers as key predictors of COVID-19 incidence, mortality, and epidemic growth rate at the country level. In addition, we used the strong association with temperature to generate a maximum likelihood estimate of the true epidemic start date, revealing that community transmission likely began on average 1–2 months prior to detection of the first reported case.

Several studies have examined weather factors (29, 30), demographic factors (12), health factors (13), geographic factors (31), and planetary factors (14, 32) in relation to COVID-19 incidence and mortality. To our knowledge, this is the first study to do so at high spatial and temporal resolution using global incidence, death, and weather data, and to simultaneously examine a panel of demographic, health, and geographic factors. Our findings strongly link epidemic growth rate, incidence, and mortality with lower temperatures (6–15°C) and lower humidity at the global scale, corroborating the findings of two studies in China (6, 30). Our results suggest that temperatures <15°C, but especially between 6 and 15°C, may be particularly conducive to COVID-19 spread, and argue for especially vigilant surveillance in countries anticipating average daily temperatures in this range over the coming year (Figure 5). Other weather variables did not offer any additional explanatory power in our analyses. Our findings are consistent with the proposed mechanism of COVID-19 transmission, and what is known about other viral infections transmitted primarily by respiratory droplet, such as influenza (33). Cold and dry climates are thought to be conducive to viral survival in the environment and host susceptibility to infection (34). Relative humidity and temperature have been shown to determine respiratory droplet evaporation kinetics and ultimate equilibrium droplet size, which in turn determines how long the droplet will remain suspended in air and where it will deposit in the respiratory system once inhaled (35). Increased temperature also promotes more rapid viral degradation (36). Although UV light has been shown to promote viral degradation in laboratory settings (37), including in SARS-CoV-1 (38), our observed association between COVID-19 and reduced sunshine appears to be entirely explained by covariation in temperature. Our findings regarding temperature and humidity may provide an explanation for outbreaks related to indoor air-conditioning systems (39), and highlight the importance of further research on indoor transmission risk factors. Our estimation of the true epidemic start date (5–7 weeks prior to first reported case) is in line with findings from retrospective screening studies (40, 41). Our analysis suggests 1–2 months of undetected COVID-19 community transmission as a ubiquitous feature of COVID-19 epidemics, a feature that almost certainly promoted rapid regional and global spread. We speculate that the 5–7 week delay in detection that we observe was likely due to combination of factors: high degree of pre-symptomatic COVID-19 transmission (42), delayed recognition of community spread (40), and delayed roll-out of testing. Our analysis demonstrates the feasibility of using a weather variable to estimate and predict the timing of community transmission in a fast-moving pandemic, when widespread testing and serologic data may not yet be available.

**Figure 5.**
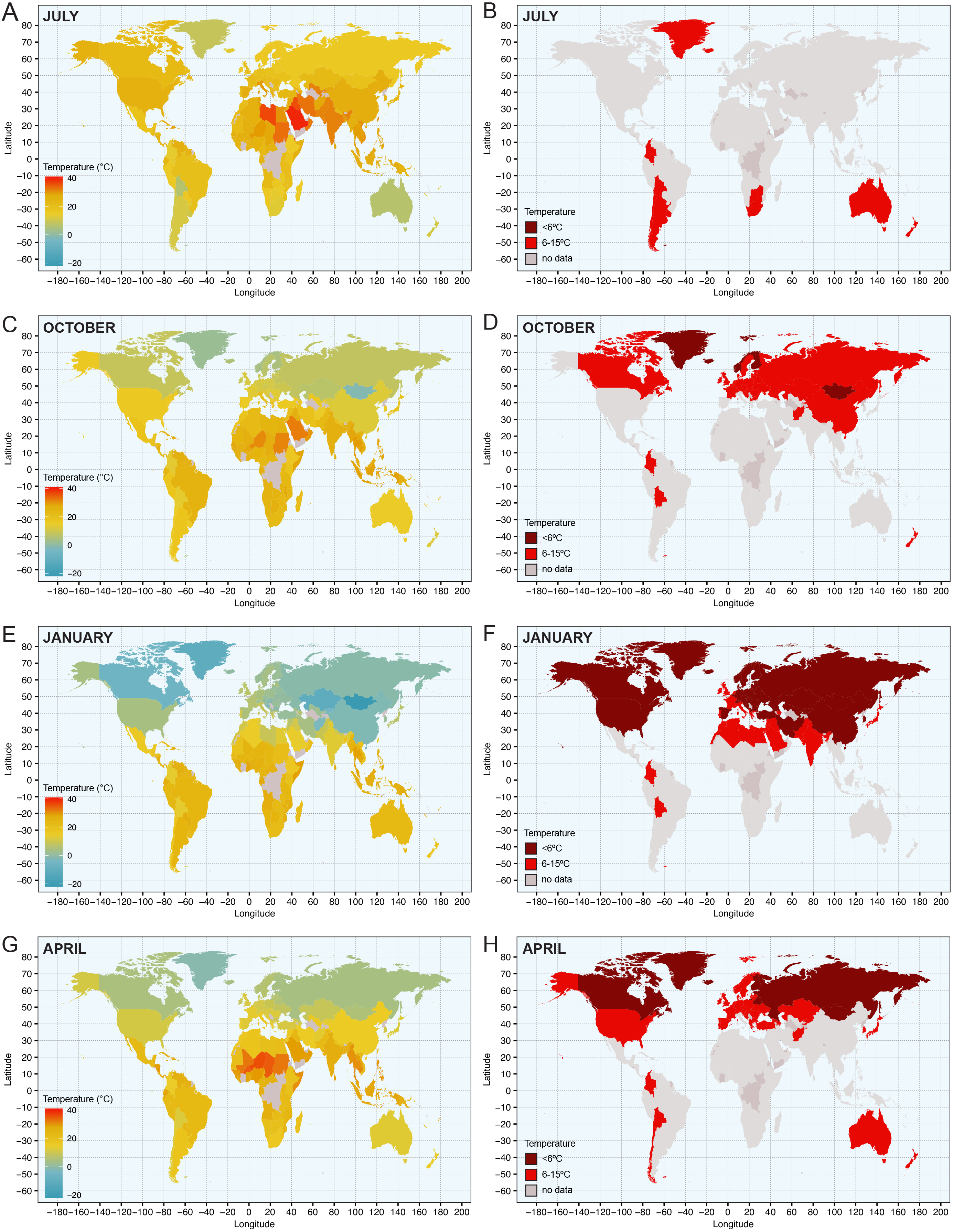
World temperature. (A-H) World maps of average monthly temperature in each country, shaded by capital city temperature, in (A and B) July 2019, (C and D) October 2019, (E and F) January 2020, and (G and H) April 2020. B, D, F, and H highlight countries that had average monthly temperatures of <6°C (dark red) or 6-15°C (red) in the corresponding month.

A number of studies have linked older age, obesity, and chronic medical conditions such as diabetes and hypertension to increased COVID-19 severity (13, 43), and it is therefore not surprising that BMI and obesity were significantly associated with COVID-19 incidence and mortality at the country level. Although we observed a strong association between age and COVID-19 outcomes in univariable analyses, age was not independently predictive in multivariable models. Prior studies have reported mixed findings on the effect of cigarette smoking on COVID-19 risk and severity (44–46). Our analysis points to a clear and significant association between increased cigarette consumption and increased national COVID-19 outcomes in univariate and odds ratio analyses, which aligns with well-established literature on increased severity of respiratory infections due to smoking (47). In multivariable modeling, however, this association did not reach statistical significance. This may be due, in part, to the fact that per capita cigarette consumption is a poor proxy for actual exposure. Our observed association with number of air travelers likely reflects a potentiating effect of repeated ‘seeding’ of COVID-19 on epidemic initiation and growth. Higher altitude was strongly associated with epidemic growth rate, even when adjusting for temperature and humidity. It is possible that the air at higher altitudes is more conducive to COVID-19 transmission for unknown reasons, and this may explain lay reports of especially explosive epidemics in ski resorts across the world. Our observed positive association with increased HDI is surprising and appears to contradict our negative association with number of hospital beds. These associations are not explained by differences in testing, as they remain significant when adjusting for total tests performed.

Our study is limited by its ecologic study design, precluding unbiased inference regarding causality or individual risk (48). In addition, although we attempted to include an exhaustive list of potentially confounding variables in this study, our analysis did not include data on all potential confounders. For instance, we could not control for differences in mitigation efforts (such as social distancing and national lockdown orders), accuracy of case and death reporting, number of superspreaders, or genetic susceptibility to infection. Use of capital city weather data also limited the resolution of our analyses, as these may not necessarily represent the weather of the entire country, particularly for countries with larger land masses. This was necessary, however, given the unavailability of accurate city-level incidence and death data for most countries. In addition, many of the demographic variables included in this analysis are measured at the population level and may therefore fail to accurately reflect risks faced at the individual level. Finally, it should be noted that the factors included in this analysis are unable to explain all of the variability in COVID-19 burden. Some generally warm and humid countries, such as Brazil and Indonesia, are experiencing severe COVID-19 epidemics. This suggests that epidemic intensity cannot be fully explained by weather or population characteristics, and mitigation strategies should be enforced. This study was strengthened by inclusion and simultaneous examination of data on a wide range of weather, demographic, health, geographic, and planetary factors. Our findings may help guide prevention and mitigation efforts by predicting future epidemic hotspots and the likelihood and timing of seasonal peaks in incidence. They also highlight the need for widespread and early testing to detect ongoing community transmission, particularly in areas anticipating the arrival of lower temperatures. Further study of the weather, sociodemographic, and geographic factors governing the rate of COVID-19 transmission will be critical to combatting this unprecedented pandemic.

## Data Availability

Data referred to in this manuscript will be made publicly available at the time of peer-reviewed publication.

## ACKNOWLEDGEMENTS

We thank Drs. Ann Chao and Marc Bulterys for their careful reading of the manuscript.

## AUTHOR CONTRIBUTIONS

N.Y.L and P.L.B designed research. N.Y.L, M.A.B., and P.L.B performed research. N.Y.L and P.L.B. analyzed data. N.Y.L, M.A.B., and P.L.B wrote the paper.

## REFERENCES

1. He J, Tao H, Yan Y, Huang SY, Xiao Y. Molecular mechanism of evolution and human infection with SARS-CoV-2. Viruses. 2020;12(4).

2. Sungnak W, Huang N, Bécavin C, et al. SARS-CoV-2 entry factors are highly expressed in nasal epithelial cells together with innate immune genes. Nature Medicine. 2020;26:681–7.

3. Zhang T, Wu Q, Zhang Z. Probable pangolin origin of SARS-CoV-2 associated with the COVID-19 Outbreak. Current Biology. 2020;30(7):1346–51.

4. van Doremalen N, Bushmaker T, Morris DH, Holbrook MG, Gamble A, Williamson BN, et al. Aerosol and Surface Stability of SARS-CoV-2 as Compared with SARS-CoV-1. N Engl J Med. 2020;382(16):1564–7.

5. Modes of transmission of virus causing COVID-19: implications for IPC precaution recommendations [press release]. WHO Scientific BriefMarch 29, 2020.

6. Sun Z, Thilakavathy K, Kumar SS, He G, Liu SV. Potential Factors Influencing Repeated SARS Outbreaks in China. International Journal of Environmental Research Public Health. 2020;17(5):1633.

7. Sajadi MM, Habibzadeh P, Vintzileos A, Shokouhi S, Miralles-Wilhelm F, Amoroso A. Temperature, humidity, and latitude analysis to predict potential spread and seasonality for COVID-19. SSRN. 2020.

8. Kissler SM, Tedijanto C, Lipsitch M, Grad Y. Social distancing strategies for curbing the COVID-19 epidemic. Lancet ID (in press) Preprint on medRxivorg. 2020 https://www.medrxiv.org/content/10.1101/2020.03.22.20041079v1.

9. Saukkoriipi A, Jokelainen J., et al. Decline in temperature and humidity increases the occurrence of influenza in cold climate. Environmental Health. 2014;13(22).

10. Liu J, Zhou J, Yao J, et al. Impact of meteorological factors on the COVID-19 transmission: A multi-city study in China. Sci Total Environ. 2020;726:138513.

11. Luo W, Majumder MS, Liu D, et al. The role of absolute humidity on transmission rates of the COVID-19 outbreak (preprint). medRxiv 2020021220022467; doi: https://doiorg/101101/2020021220022467. 2020.

12. Sun K, Chen J, Viboud C. Early epidemiological analysis of the coronavirus disease 2019 outbreak based on crowdsourced data: a population-level observational study. The Lancet Digital Health. 2020;2(4):E201–E8.

13. Yao Q, Wang P, Wang X, et al. Retrospective study of risk factors for severe SARS-CoV-2 infectious in hospitalized adult patients. Pol Arch Intern Med. 2020.

14. Conticini E, Frediani B, Caro D. Can atmospheric pollution be considered a co-factor in extremely high level of SARS-CoV-2 lethality in Northern Italy? Environmental Pollution. 2020;261.

15. Vardavas CI, Nikitara K. COVID-19 and smoking: A systematic review of the evidence. Tobacco Induced Diseases. 2020;18(March):20. doi:10.18332/tid/119324.

16. European Centre for Disease Prevention and Control. Coronavirus disease (COVID-19). https://www.ecdc.europa.eu/en 2020 [

17. Central Intelligence Agency (CIA). The World Factbook. https://www.cia.gov/library/publications/the-world-factbook/ 2020 [

18. The World Bank. The World Bank Open Data. https://data.worldbank.org 2020 [

19. International Monetary Fund (IMF). IMF Data. https://www.imf.org/en/Data 2020 [

20. The United Nations Human Development Programme. Human Development Reports and Data. http://hdr.undp.org/en/data 2020 [

21. World Health Organization (WHO). The Global Health Observatory: Explore a world of health data. https://www.who.int/data/gho 2020 [

22. The Tobacco Atlas. The Tobacco Atlas. https://tobaccoatlas.org 2020 [

23. Health Effects Institute (HEI). What HEI does to improve air quality and public health. https://www.healtheffects.org 2020 [

24. Germanwatch. Global Climate Risk Index 2020. https://germanwatch.org/en/17307 2020 [

25. https://TuTiempo.net. World weather and climate. https://www.tutiempo.net 2020 [

26. Bulterys PL, Bulterys MA, Phommasone K, Luangraj M, Mayxay M, Kloprogge S, et al. Climatic drivers of melioidosis in Laos and Cambodia: a 16-year case series analysis. The Lancet Planetary Health. 2018;2(8):E334–E43.

27. Bulterys PL, Le T, Quang VM, Nelson KE, Lloyd-Smith JO. Environmental predictors and incubation period of AIDS-associated penicillium marneffei infection in Ho Chi Minh City, Vietnam. Clin Infect Dis. 2013;56(9):1273–9.

28. BM B. Princeton: Princeton University Press; Ecological models and data in R 2008.

29. Al-Rousan N, Al-Najjar H. The correlation between the spread of COVID-19 infections and weather variables in 30 Chinese provinces and the impact of Chinese government mitigation plans. Eur Rev Med Pharmacol Sci. 2020;24(8):4565–71.

30. Wang J, Tang K, Feng K, Lv W. High temperature and high humidity reduce the transmission of COVID-19. SSRN. 2020.

31. Briz-Redón Á, Serrano-Aroca Á. A spatio-temporal analysis for exploring the effect of temperature on COVID-19 early evolution in Spain. Sci Total Environ. 2020;728:138811.

32. Liu K, Zhou J, Yao J, et al. Impact of meteorological factors on the COVID-19 transmission: A multi-city study in China. Sci Total Environ. 2020;726:138513.

33. Moriyama M, Hugentobler WJ, Iwasaki A. Seasonality of Respiratory Viral Infections. Annual Review of Virology. 2020;7:2.1–2.19.

34. Dowell SF. Seasonal variation in host susceptibility and cycles of certain infectious diseases. Emerging Infectious Diseases. 2001;7(3):369–74.

35. Marr LC, Tang JW, Van Mullekom J, Lakdawala SS. Mechanistic insights into the effect of humidity on airborne influenza virus survival, transmission and incidence. Journal of the Royal Society Interface. 2019;16(150).

36. Chin AWH, Chu JTS, Perera MRA, Hui K, et al. Stability of SARS-CoV-2 in different environmental conditions. The Lancet Microbe (In Press, corrected proof) https://doiorg/101016/S2666-5247(20)30003-3. 2020.

37. Lytle DC, Sagripanti JL. Predicted inactivation of viruses of relevance to biodefense by solar radiation. Journal of Virology. 2005;79(22):14244–52.

38. Duan SM, Zhao XS, Wen RF, et al. Stability of SARS coronavirus in human specimens and environment and its sensitivity to heating and UV irradiation. Biomed Environ Sci. 2003;16(3):246–55.

39. Lu J, Gu J, Li K, et al. COVID-19 outbreak associated with air conditioning in restaurant, Guangzhou, China, 2020. Emerging Infectious Diseases. 2020;26(7).

40. Hogan CA, Sahoo MK, Pinsky BA. Sample Pooling as a Strategy to Detect Community Transmission of SARS-CoV-2. JAMA. 2020.

41. Deslandes A, Berti V, Tandjaoui-Lambotte Y, Alloui C, Carbonnelle E, Zahar JR, et al. SARS-COV-2 was already spreading in France in late December 2019. Int J Antimicrob Agents. 2020:106006.

42. Arons MM, Hatfield KM, Reddy SC, Kimball A, James A, Jacobs JR, et al. Presymptomatic SARS-CoV-2 Infections and Transmission in a Skilled Nursing Facility. N Engl J Med. 2020.

43. Zhang JJ, Xiang D, Cao Y, et al. Clinical characteristics of 140 patients infected with SARSCoV-2 in Wuhan, China. Allergy. 2020.

44. Farsalinos K, Barbouni A, Niaura R. Smoking, vaping and hospitalization for COVID-19. Pre-print on Qeioscom https://doiorg/1032388/Z69O8A13. 2020.

45. Propper RE. Does cigarette smoking protect against SARS-CoV-2 infection? Nicotine and Tobacco Research. 2020.

46. Patanavanich R, Glantz SA. Smoking is Associated with COVID-19 Progression: A Meta-Analysis. Nicotine Tob Res. 2020.

47. U.S. Department of Health and Human Services, Centers for Disease Control and Prevention, National Center for Chronic Disease Prevention and Health Promotion, Office on Smoking and Health. The health consequences of smoking: 50 years of progress – A report by the Surgeon General, Atlanta. 2014.

48. Morgenstern H. Ecologic studies in epidemiology: concepts, principles, and methods. Annu Rev Public Health. 1995;16:61–81.

